# Sex Differences in the Clinical and Imaging Characteristics of Korean CADASIL Patients: A Korean Nationwide Retrospective Study

**DOI:** 10.1101/2025.04.28.25326613

**Authors:** Joong-Goo Kim, Jay Chol Choi, Chul-Hoo Kang, Jung-Hwan Oh, Jung Seok Lee, Ji-Hoon Kang, Sukyung Byeon, Jae-Sung Lim, Bum Joon Kim, Hyunjin Kim, Chong Hyun Suh, Sung Hyuk Heo, Ho Geol Woo, Hyo Suk Nam, Byung-Chul Lee, Kyung-Ho Yu, Mi Sun Oh, Minwoo Lee, Chi Kyung Kim, Kyungmi Oh, Sung Hoon Kang, Keon-Joo Lee, Jung Hoon Han, Young Seo Kim, Hyun Young Kim, Hee-Jin Kim, Hee-Joon Bae, Moon-Ku Han, Jihoon Kang, Beom Joon Kim, Jun Yup Kim, Seung-Hoon Lee, Keun-Hwa Jung, Sung-Il Sohn, Jeong-Ho Hong, Hyungjong Park, Jee Hyun Kwon, Wook-Joo Kim, Dong-Ick Shin, Kyu Sun Yum, Hee-Yun Chae, Sang-Min Sung, Sang Won Seo, Jun Pyo Kim, Jin-Man Jung, Kyung Bok Lee, Tai Hwan Park, Sang-Soon Park, Jin Kyo Choi, Man-Seok Park, Joon-Tae Kim, Kang-Ho Choi, Jae-Kwan Cha, Dae-Hyun Kim, Byeol-A Yoon, Soo Joo Lee, Jae Guk Kim, Do-Hyung Kim, Dong-Eog Kim, Dong-Seok Gwak, Chul-Ho Kim, Sang-Hwa Lee, Jun Lee, Doo Hyuk Kwon, Kyusik Kang, Kwang-Yeol Park, Hae-Bong Jeong, Chan-Young Park, Yong-Jin Cho, Keun-Sik Hong, Hong-Kyun Park

## Abstract

**Background:** Cerebral autosomal dominant arteriopathy with subcortical infarcts and leukoencephalopathy (CADASIL) is a hereditary small vessel disease caused by *NOTCH3* mutations. While CADASIL affects both sexes, differences in clinical presentation and brain magnetic resonance imaging (MRI) findings remain unclear. This study aims to investigate sex-specific variations in vascular risk factors, clinical manifestations, and brain MRI features among Korean CADASIL patients.

**Methods:** A nationwide retrospective study analyzed 368 CADASIL patients (179 men, 189 women). Clinical characteristics, vascular risk factors, and *NOTCH3* variants were compared between sexes. Brain MRI findings, including white matter hyperintensity, lacunes, cerebral microbleeds, atrophy, and enlarged perivascular spaces, were evaluated. Kaplan-Meier analysis assessed sex differences in the onset of ischemic stroke, intracerebral hemorrhage (ICH), and dementia, and recurrent headaches.

**Results:** Men exhibited a higher prevalence of ischemic stroke (70.9% vs. 44.4%, p<0.001), whereas women were more likely to report recurrent headaches (35.4% vs. 16.9%, p<0.001). Survival analysis showed that men had a significantly earlier onset of ischemic stroke (Hazard Ratio[HR]: 2.78, 95% CI: 1.92-4.03, p<0.001) while being male was associated with significantly later onset of recurrent headache (HR: 0.59, 95% CI: 0.38-0.90, p=0.016). No significant sex differences were observed for ICH (HR: 2.04, 95% CI: 0.80-5.18, p=0.136) or dementia (HR: 1.26, 95% CI: 0.56-2.84, p=0.579). On brain MRI, male sex was associated with significantly higher risk for lacune burden (Odds Ratio[OR] 3.03, 95% CI 1.70-5.38, p<0.001) along with age (OR 1.03, 95% CI 1.01-1.05, p=0.002) and hypertension (OR 1.86, 95% CI 1.15-3.03, p=0.002).

**Conclusion:** This study highlights significant sex-based differences in CADASIL, with men experiencing earlier and higher risk of ischemic stroke, along with a greater burden of lacunes, while women showed an increased likelihood of recurrent headaches at an earlier age. These findings underscore the importance of integrating sex-specific considerations into CADASIL prognosis and treatment planning.

## Introduction

Cerebral autosomal dominant arteriopathy with subcortical infarcts and leukoencephalopathy (CADASIL) is the most common monogenic form of small vessel disease, caused by mutations in the *NOTCH3* gene.^1,2^ The disease is characterized by recurrent strokes, cognitive decline, migraine, and psychiatric symptoms.^3^ While CADASIL is widely studied, increasing evidence suggests that sex plays a crucial role in its clinical presentation and progression.^4-10^

Previous studies have indicated that male CADASIL patients tend to experience a more aggressive disease course, with higher risk of stroke, dementia, disability, or shorter survival compared to female patients.^4,6,8-10^ For instance, a retrospective study of 411 CADASIL patients found that men had a significantly lower median age at death (64.6 years) compared to women (70.7 years), and they also became bedridden at an earlier age.^8^ Additionally, male patients had an approximately two-fold increased risk of stroke and dementia compared with female patients after adjustment for age, *NOTCH3* variant position, and other vascular risk factors.^4,9^ Interestingly, the age of onset of stroke did not show a significant difference between men and women although a trend toward an earlier onset of stroke was seen in males.^5,6,8^ Conversely, female patients showed a higher prevalence of migraine with aura.^5^

Regarding brain magnetic resonance imaging (MRI), lacune number or volumes were significantly greater in male patients than in female patients.^4,6^ Males also tend to have a higher number of cerebral microbleeds (CMBs) and more pronounced brain atrophy. Neuroimaging studies using 7-Tesla MRI have further highlighted sex-specific differences in brain structure. Male CADASIL patients exhibit greater frontotemporal atrophy, particularly in the orbitofrontal and anterior cingulate cortices, regions associated with executive dysfunction.^7^ This aligns with findings that men suffer from more severe executive dysfunction despite similar global cognitive scores between the sexes.^5^

Despite these observations, a comprehensive understanding of sex differences in CADASIL remains elusive, particularly in Asian populations, where clinical manifestations and genetic variants are different from those in the above-mentioned European CADASIL cohorts; later onset, more frequent occurrence of CMBs and intracerebral hemorrhage (ICH), and distinct *NOTCH3* variants (R75P and R544C).^11-13^ Therefore, this study aims to elucidate the sex-specific variations in clinical manifestations and neuroimaging characteristics of Korean CADASIL patients.

## Methods

Korean nationwide retrospective CADASIL study enrolled patients aged 19 years or older who had been either confirmed or suspected of having CADASIL through genetic testing from 26 nationwide institutions in South Korea (Figure 1). Eligible participants included individuals identified with a pathogenic variant, likely pathogenic variant, or a variant of unknown significance. The study period covered patients diagnosed with or suspected of CADASIL through genetic testing from January 1, 2013, to December 31, 2023. We collected data from medical records until July 31, 2024. Patients whose clinical, genetic, or imaging data could not be sufficiently assessed through medical records were excluded from the study. The nationwide study was able to collect data from 445 Korean CADASIL patients who were confirmed or suspected through genetic testing. Of the 445 patients, 368 (82.7%) were confirmed to have pathogenic variants, 46 (10.3%) had variants of undetermined significance, and 31 (7.0%) had benign variants based on the ClinVar database (https://www.ncbi.nlm.nih.gov/clinvar/) as of November 30, 2024. Of 445 patients, the current study analyzed clinical, genetic, and neuroradiological findings from the 368 patients with pathogenic *NOTCH3* variants. This study was approved by the Institutional Review Board (IRB) of Jeju National University Hospital (IRB No. 2023-08-012) and the respective IRBs of all participating institutions prior to initiating the study.

**Figure 1.**
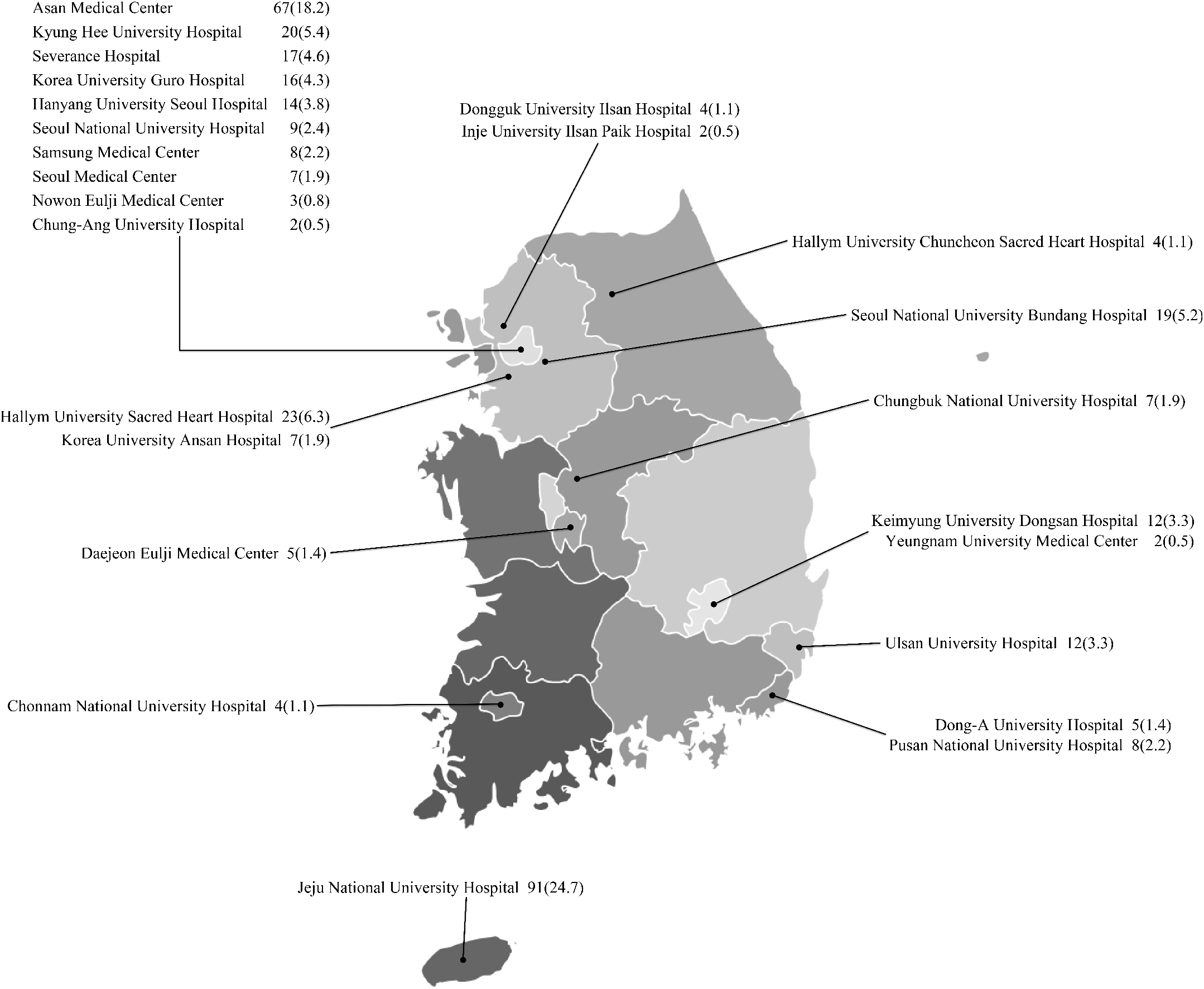
Contributing centers and number of patients in each center

The collected data encompassed 1) demographic information such as age, sex, height, weight, occupation, and educational background, as well as family history, including stroke, dementia, depression, and migraine, 2) genetic test results detailing the location of genetic variants and amino acid changes, 3) clinical symptoms and their onset age, such as stroke (ischemic, hemorrhagic), cognitive impairment, dementia, recurrent headaches, parkinsonism and epilepsy, were also recorded, 4) vascular risk factors—including hypertension, diabetes, hyperlipidemia, smoking, alcohol intake, atrial fibrillation, 5) information on medications for hypertension, hyperlipidemia, diabetes, antithrombotic therapy, and dementia treatment, 6) brain MRI were reviewed, along with laboratory assessments such as blood tests, and blood pressure, pulse rate, and electrocardiogram results, and 7) cognitive and disability tests results, including modified Rankin Scale (mRS) score^14^, Korean Mini-Mental State Examination(K-MMSE)^15^, and the Seoul Neuropsychological Screening Battery (SNSB).^16,17^ We used the raw K-MMSE score and z-scores for five cognitive domains (attention, language, visuospatial functions, memory and frontal/executive functions from the SNSB test for the analyses. In this study, recurrent headaches were included as one of the manifestations of CADASIL symptoms rather than being classified as migraine, because previous studies have reported that migraine was found in only a minority of Korean CADASIL patients with headaches, and other headaches were classified as tension-type or unclassifiable, depending on the genotype.^12,18^

In this study, the collection and analysis of brain MRI data were led by JS Lim and CH Suh from Asan Medical Center. Participating institutions anonymized all DICOM headers before transmitting MRI data. MRI analysis was conducted in accordance with the STRIVE (Standards for Reporting Vascular Change Neuroimaging) criteria.^19^ Brain MRI scans were rated for white matter hyperintensities (WMH), lacune, cerebral microbleed (CMB), dilated perivascular space (dPVS), and brain atrophy according to the CADA-MRIT criteria suggested by Zhang et al.^20^ All ratings were performed by the same neuroradiologist (CH Suh, 14 years of clinical experience in neuroradiology). We also assessed the presence of WMH on anterior temporal lobe and external capsule which are frequently seen in patients with CADASIL.^21^ Additionally, brain volume and WMH volume were measured by using commercialized, automated software using deep learning (VUNO Med-DeepBrain, Seoul, South Korea), and normalized WMH volume (nWMHv) was calculated.^22,23^

In this retrospective study, we collected all brain MRI scans conducted during the study period. Therefore, many patients had multiple brain MRI scans. From these MRI scans, we first selected the scans closest to the diagnosis date and that contained the greatest number of sequences to assess WMH, lacune, CMBs, and brain atrophy. Second, we selected brain MRI scans that best allowed for the evaluation of WMH, lacune, CMBs, or brain atrophy, even if they were examined on different dates. The second MRI selection was used for sensitivity analysis.

### Statistical analysis

Continuous variables were summarized as means with standard deviations and compared between sexes using independent t-tests or Mann-Whitney U tests, as appropriate. Categorical variables were presented as frequencies with percentages and analyzed using chi-square or Fisher’s exact tests. Multivariable logistic regression was performed to assess associations between sex and clinical manifestations including ischemic stroke, ICH, and dementia, adjusting for age, vascular risk factors, and *NOTCH3* variant position. For recurrent headaches, only age and *NOTCH3* variant position were adjusted. Kaplan-Meier survival analysis with log-rank tests evaluated sex differences in the onset of ischemic stroke, ICH, recurrent headaches and dementia. In this retrospective study, patients lost to follow-up were considered censored at their last follow-up date. Cox proportional hazards models were applied to estimate hazard ratios for these outcomes. Ordinal logistic regression was used to examine the associations between age, vascular risk factors, and *NOTCH3* variant position with markers of cerebral small vessel diseases, except for nWMHv, which was analyzed using linear regression. Sensitivity analyses were conducted using the second MRI dataset to confirm robustness. A significance level of p<0.05 was considered statistically significant. All statistical analyses were performed using the Stata data analysis software (Version 18, StataCorp, College Station, Texas, USA).

## Results

### Demographic characteristics

The mean age of the patients was 61.7±11.9 years, and 48.6% were male. Among vascular risk factors, hypertension was found in 44.0% of patients, diabetes in 19.0%, hypercholesterolemia in 48.2%, smoking in 30.0%, and alcohol consumption in 29.2%. Men (N=179) were significantly younger than women (N=189) (59.1 ± 11.2 vs. 64.2 ± 12.1 years, p<0.001). Hypertension (45.3% vs. 42.9%, p=0.644) and hypercholesterolemia (48.6% vs. 47.9%, p=0.889) were similarly prevalent in both sexes. However, men had a significantly higher prevalence of diabetes mellitus (25.7% vs. 12.7%, p=0.001), smoking (56.1% vs. 3.6%, p<0.001), and alcohol drinking (45.2% vs. 13.3%, p<0.001). In terms of medication use, antihypertensive agents, antiplatelets, statins, and acetylcholinesterase inhibitors were prescribed to 45.9%, 83.4%, 75.8%, and 19.8% of patients, respectively (Table 1). The median follow-up duration was 45 months (interquartile range, 17–83 months).

**Table 1.**
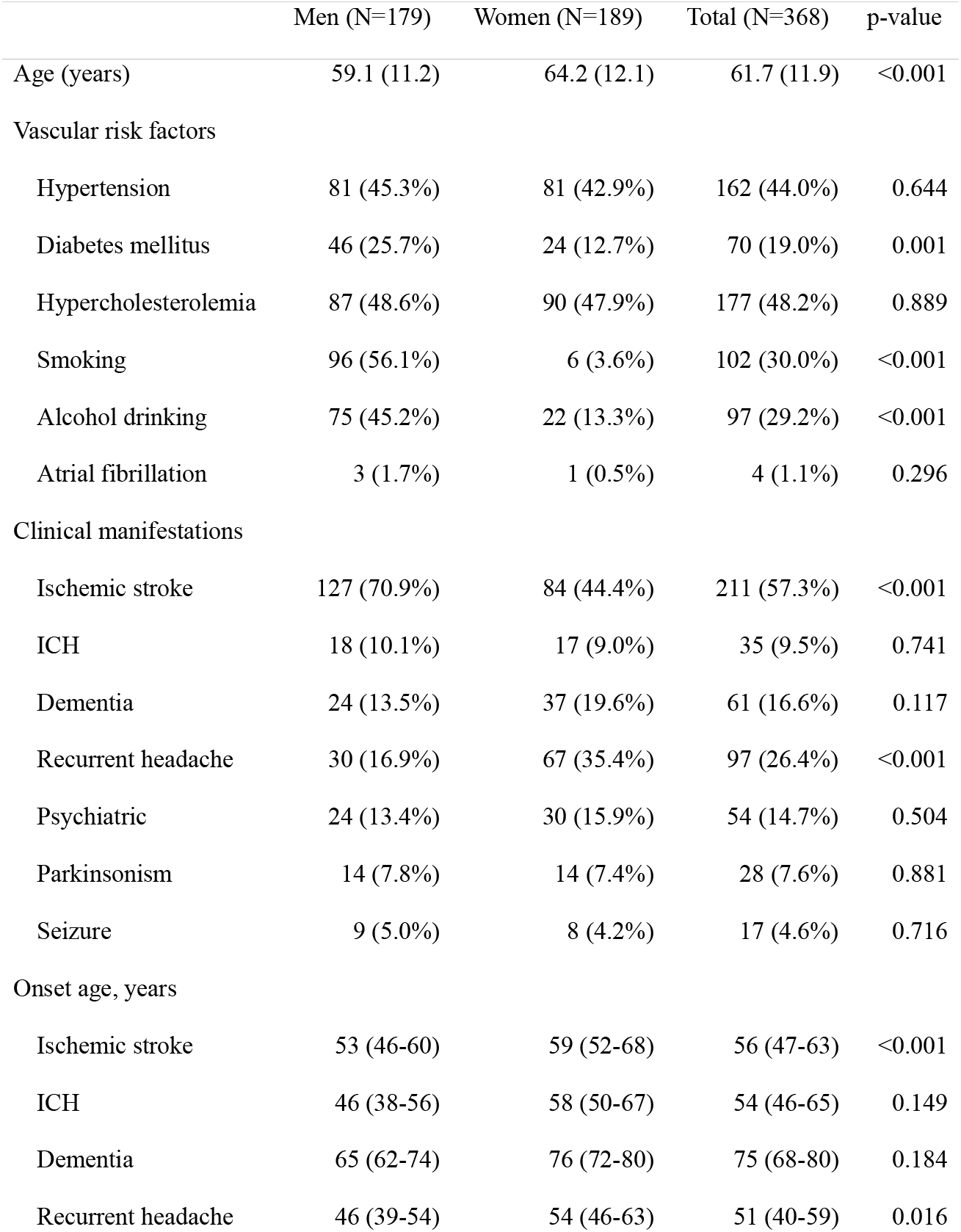
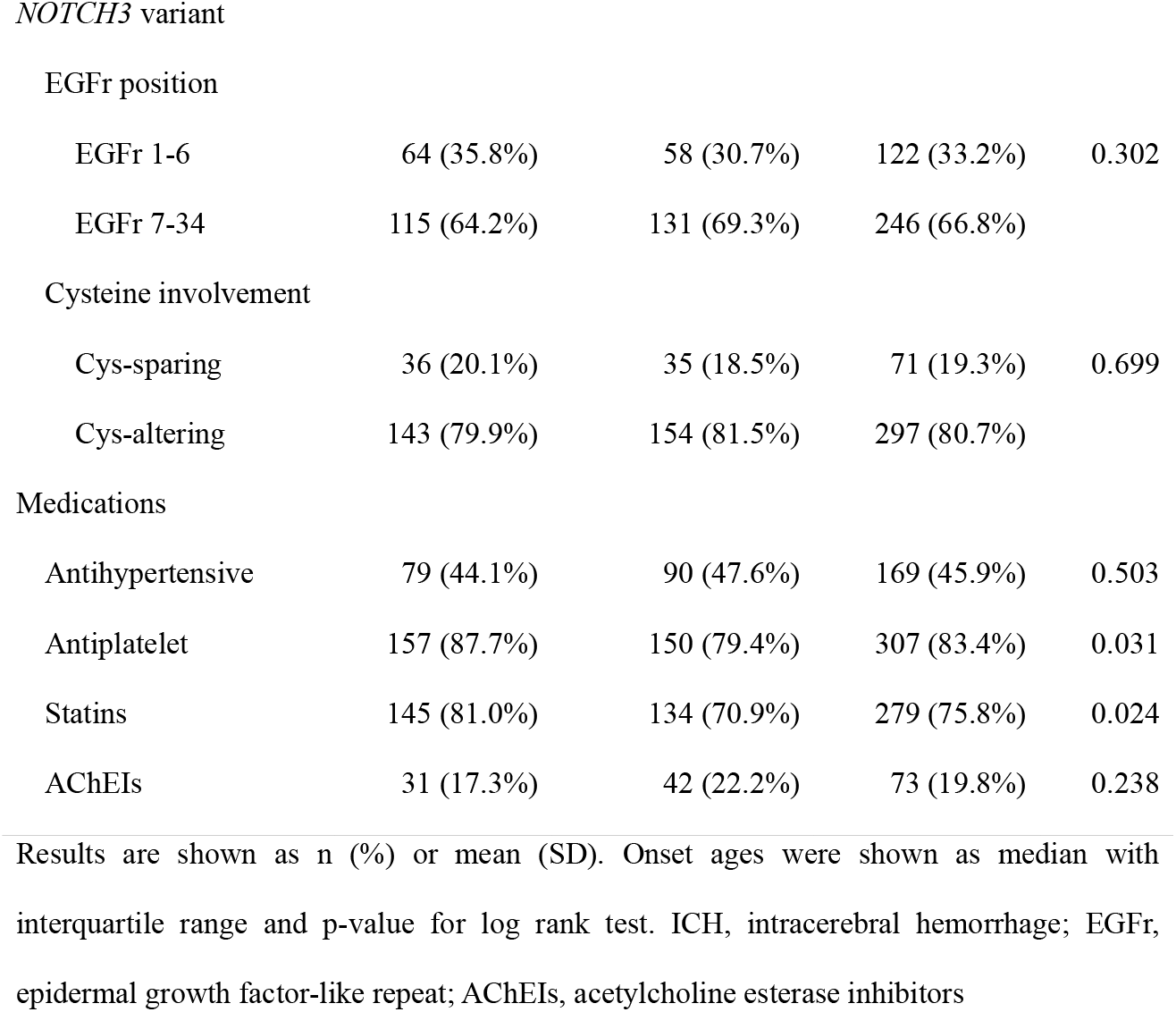
Characteristics of the patients by sex.

|  | Men (N=179) | Women (N=189) | Total (N=368) | p-value |
| --- | --- | --- | --- | --- |
| Age (years) | 59.1 (11.2) | 64.2 (12.1) | 61.7 (11.9) | <0.001 |
| Vascular risk factors |  |  |  |  |
| Hypertension | 81 (45.3%) | 81 (42.9%) | 162 (44.0%) | 0.644 |
| Diabetes mellitus | 46 (25.7%) | 24 (12.7%) | 70 (19.0%) | 0.001 |
| Hypercholesterolemia | 87 (48.6%) | 90 (47.9%) | 177 (48.2%) | 0.889 |
| Smoking | 96 (56.1%) | 6 (3.6%) | 102 (30.0%) | <0.001 |
| Alcohol drinking | 75 (45.2%) | 22 (13.3%) | 97 (29.2%) | <0.001 |
| Atrial fibrillation | 3 (1.7%) | 1 (0.5%) | 4 (1.1%) | 0.296 |
| Clinical manifestations |  |  |  |  |
| Ischemic stroke | 127 (70.9%) | 84 (44.4%) | 211 (57.3%) | <0.001 |
| ICH | 18 (10.1%) | 17 (9.0%) | 35 (9.5%) | 0.741 |
| Dementia | 24 (13.5%) | 37 (19.6%) | 61 (16.6%) | 0.117 |
| Recurrent headache | 30 (16.9%) | 67 (35.4%) | 97 (26.4%) | <0.001 |
| Psychiatric | 24 (13.4%) | 30 (15.9%) | 54 (14.7%) | 0.504 |
| Parkinsonism | 14 (7.8%) | 14 (7.4%) | 28 (7.6%) | 0.881 |
| Seizure | 9 (5.0%) | 8 (4.2%) | 17 (4.6%) | 0.716 |
| Onset age, years |  |  |  |  |
| Ischemic stroke | 53 (46-60) | 59 (52-68) | 56 (47-63) | <0.001 |
| ICH | 46 (38-56) | 58 (50-67) | 54 (46-65) | 0.149 |
| Dementia | 65 (62-74) | 76 (72-80) | 75 (68-80) | 0.184 |
| Recurrent headache | 46 (39-54) | 54 (46-63) | 51 (40-59) | 0.016 |
| EGFr 1-6 | 64 (35.8%) | 58 (30.7%) | 122 (33.2%) | 0.302 |
| EGFr 7-34 | 115 (64.2%) | 131 (69.3%) | 246 (66.8%) |  |
| Cysteine involvement |  |  |  |  |
| Cys-sparing | 36 (20.1%) | 35 (18.5%) | 71 (19.3%) | 0.699 |
| Cys-altering | 143 (79.9%) | 154 (81.5%) | 297 (80.7%) |  |
| Medications |  |  |  |  |
| Antihypertensive | 79 (44.1%) | 90 (47.6%) | 169 (45.9%) | 0.503 |
| Antiplatelet | 157 (87.7%) | 150 (79.4%) | 307 (83.4%) | 0.031 |
| Statins | 145 (81.0%) | 134 (70.9%) | 279 (75.8%) | 0.024 |
| AChEIs | 31 (17.3%) | 42 (22.2%) | 73 (19.8%) | 0.238 |

Results are shown as n (%) or mean (SD). Onset ages were shown as median with interquartile range and p-value for log rank test. ICH, intracerebral hemorrhage; EGFr, epidermal growth factor-like repeat; AChEIs, acetylcholine esterase inhibitors

### Genetics

Regarding genetics, patients had 36 different pathogenic variants, with more than half located in exon 11 (55.2%) (Figure 2 and Table S1). The most frequent variants were R544C (40.8%), R75P (17.9%), C542R (5.4%), and R587C (4.6%). Cysteine-altering variants were identified in 80.7% of patients, while only one-third had variants located in EGFr1-6 (33.2%). There were no significant differences in the distribution of *NOTCH3* variant positions between men and women. EGFr1-6 mutations were present in 35.8% of men and 30.7% of women (p=0.302). Cysteine-altering mutations were similarly prevalent in both sexes (79.9% vs. 81.5%, p=0.699).

**Figure 2.**
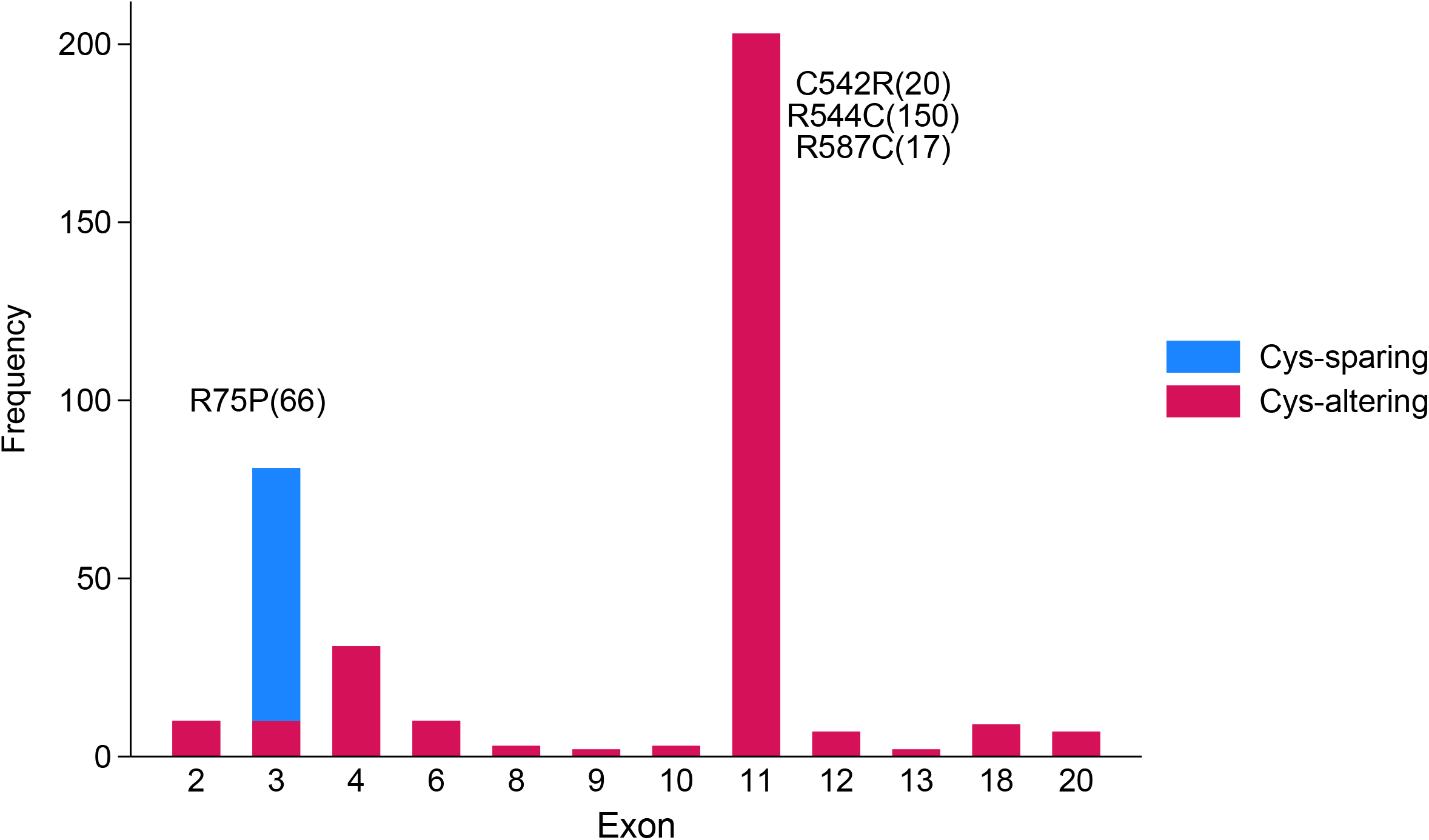
Distribution of *NOTCH3* variants by exon.

### Clinical manifestations

Ischemic stroke including transient ischemic attack was the most common clinical manifestation, occurring in 57.3% of patients, followed by recurrent headaches (26.4%), dementia (16.6%), psychiatric symptoms (14.7%), ICH (9.5%), parkinsonism (7.6%), and seizures (4.6%). Men exhibited a higher risk of ischemic stroke (70.9% vs. 44.4%, p<0.001), whereas women were more likely to report recurrent headaches (35.4% vs. 16.9%, p<0.001). Dementia was observed more frequently in women (19.6% vs. 13.4%), though the difference was not statistically significant (p=0.117). Male sex (Odds ratio [OR]: 4.01, 95% CI: 2.16-7.43, p<0.001), remained significantly associated with increased risk of ischemic stroke along with variant position on EGFr1-6 (OR: 2.11, 95% CI: 1.22-3.66, p=0.007), and hypertension (OR: 1.86, 95% CI: 1.10-3.16, p=0.021) on multivariable analysis. Male sex (OR: 0.32, 95% CI: 0.19-0.53, p<0.001) was associated with significantly decreased risk of having recurrent headache on multivariable analysis (Figure 3). When we stratified the regression models by hypertension or smoking status, the ORs for ischemic stroke with male sex increased further in the subgroup without hypertension (OR: 5.08, 95% CI: 2.31-11.20, p<0.001) compared with the subgroup with hypertension (OR: 3.00, 95% CI: 1.01-8.94, p=0.048) and also in the subgroup without smoking (OR: 4.83, 95% CI: 2.42-9.62, p<0.001) compared with the subgroup with smoking (OR: 1.70, 95% CI: 0.27-10.66, p=0.573) suggesting independent association between male sex and increased risk of ischemic stroke regardless of hypertension or smoking (Figure S1).

**Figure 3.**
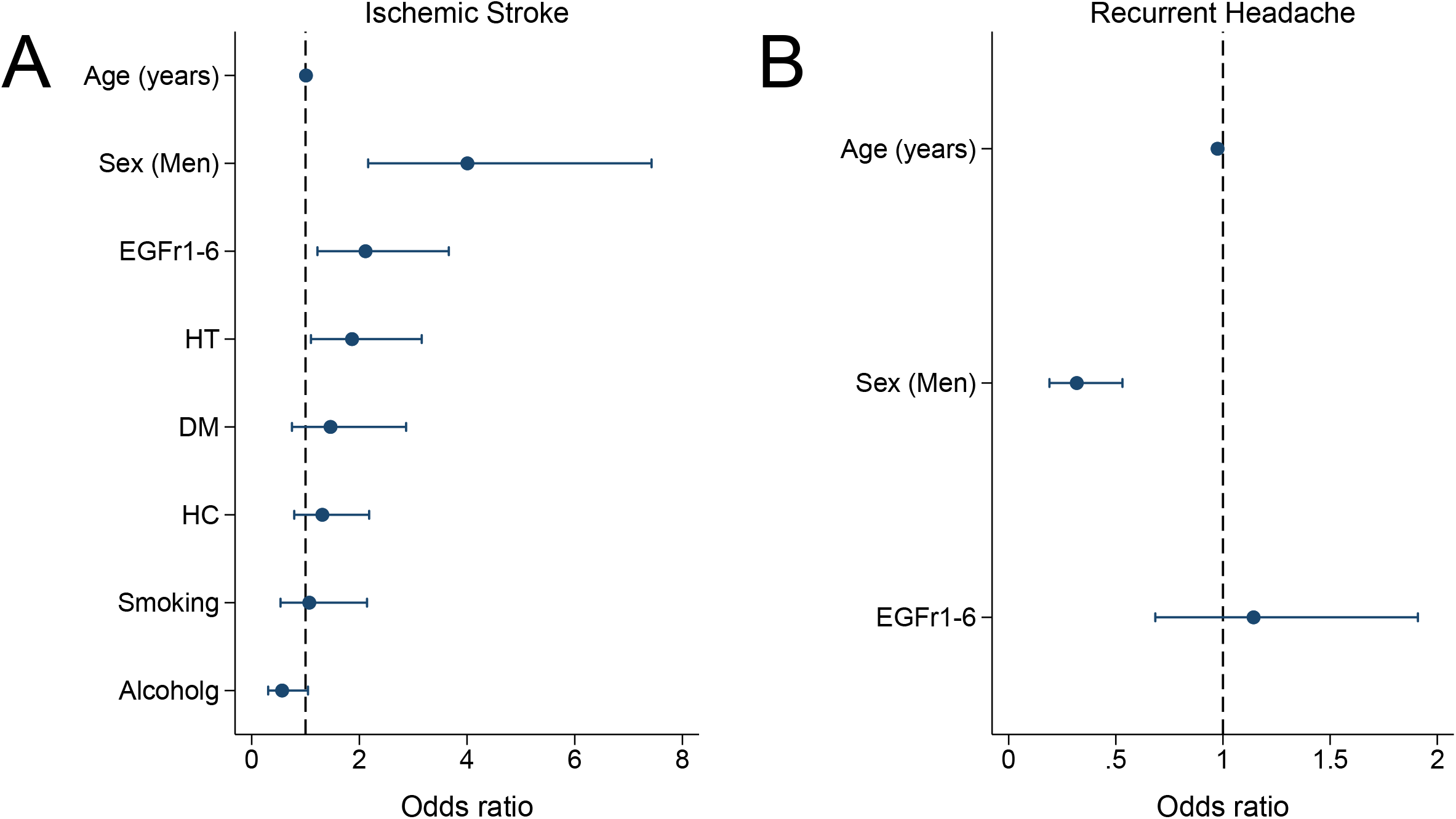
Factors associated with ischemic stroke and recurrent headache on multivariable analyses. HT, hypertension; DM, diabetes mellitus; HC, hypercholesterolemia

### Age of onset of clinical symptoms

The median age of onset for ischemic stroke was 56 (interquartile ranged from 47 to 63) years, while the median ages of onset for ICH, recurrent headaches, and dementia were 54 (46-65), 75 (68-80), and 51 (40-59) years, respectively. Male sex showed significantly different survival free of ischemic stroke or recurrent headache compared with female sex on log rank test (p<0.001 for ischemic stroke, p=0.016 for recurrent headache). Male sex (HR: 2.78, 95% CI: 1.92-4.03, p<0.001) and EGFr1-6 (HR: 2.09, 95% CI: 1.51-2.89, p<0.001) were significantly associated with earlier onset of ischemic stroke on Cox proportional hazard analysis while male sex was associated with significantly later onset of recurrent headache (HR: 0.59, 95% CI: 0.38-0.90, p=0.016) (Table 2). In subgroup analysis by hypertension status, male sex showed higher risk for earlier onset of stroke in the subgroup without hypertension (HR: 3.06, 95% CI: 1.90-4.94, p<0.001) than in the subgroup with hypertension (HR: 2.55, 95% CI: 1.39-4.69, p=0.003) suggesting independent association between earlier onset of ischemic stroke and male sex (Figure S2). No significant sex differences were observed for ICH (HR: 2.03, 95% CI: 0.80-5.18, p=0.136) or dementia (HR: 1.26, 95% CI: 0.56-2.84, p=0.579) (Table 2 and Figure 4).

**Table 2.** Factors affecting onset of ischemic stroke, intracerebral hemorrhage, dementia and recurrent headache.

|  | Ischemic stroke |  |  | ICH |  |  | Dementia |  |  | Recurrent headache |  |  |
| --- | --- | --- | --- | --- | --- | --- | --- | --- | --- | --- | --- | --- |
|  | Hazard | 95% | p-value | Hazard | 95% | p-value | Hazard | 95% | p-value | Hazard | 95% | p-value |
|  | Ratio | CI |  | Ratio | CI |  | Ratio | CI |  | Ratio | CI |  |
| Male sex | 2.78 | 1.92- | <0.001 | 2.03 | 0.80- | 0.136 | 1.26 | 0.56- | 0.579 | 0.59 | 0.38- | 0.016 |
|  |  | 4.03 |  |  | 5.18 |  |  | 2.84 |  |  | 0.90 |  |
| EGFr1-6 | 2.09 | 1.51- | <0.001 | 1.40 | 0.60- | 0.437 | 0.96 | 0.44- | 0.909 | 1.50 | 0.99- | 0.058 |
|  |  | 2.89 |  |  | 3.26 |  |  | 2.07 |  |  | 2.28 |  |
| Hypertension | 1.30 | 0.94- | 0.108 | 1.92 | 0.86- | 0.112 | 0.95 | 0.51- | 0.867 | — | — | — |
|  |  | 1.79 |  |  | 4.28 |  |  | 1.76 |  |  |  |  |
| Diabetes | 0.97 | 0.67- | 0.877 | 0.42 | 0.14- | 0.134 | 0.98 | 0.47- | 0.965 | — | — | — |
|  |  | 1.41 |  |  | 1.30 |  |  | 2.07 |  |  |  |  |
| Hypercholesterolemia | 0.91 | 0.67- | 0.567 | 1.15 | 0.54- | 0.721 | 1.11 | 0.60- | 0.750 | — | — | — |
|  |  | 1.24 |  |  | 2.45 |  |  | 2.05 |  |  |  |  |
| Smoking | 0.90 | 0.62- | 0.556 | 0.64 | 0.24- | 0.367 | 1.02 | 0.42- | 0.958 | — | — | — |
|  |  | 1.29 |  |  | 1.69 |  |  | 2.46 |  |  |  |  |
| Alcohol drinking | 1.00 | 0.71- | 0.996 | 1.59 | 0.67- | 0.292 | 0.97 | 0.44- | 0.950 | — | — | — |
|  |  | 1.41 |  |  | 3.78 |  |  | 2.18 |  |  |  |  |

**Figure 4.**
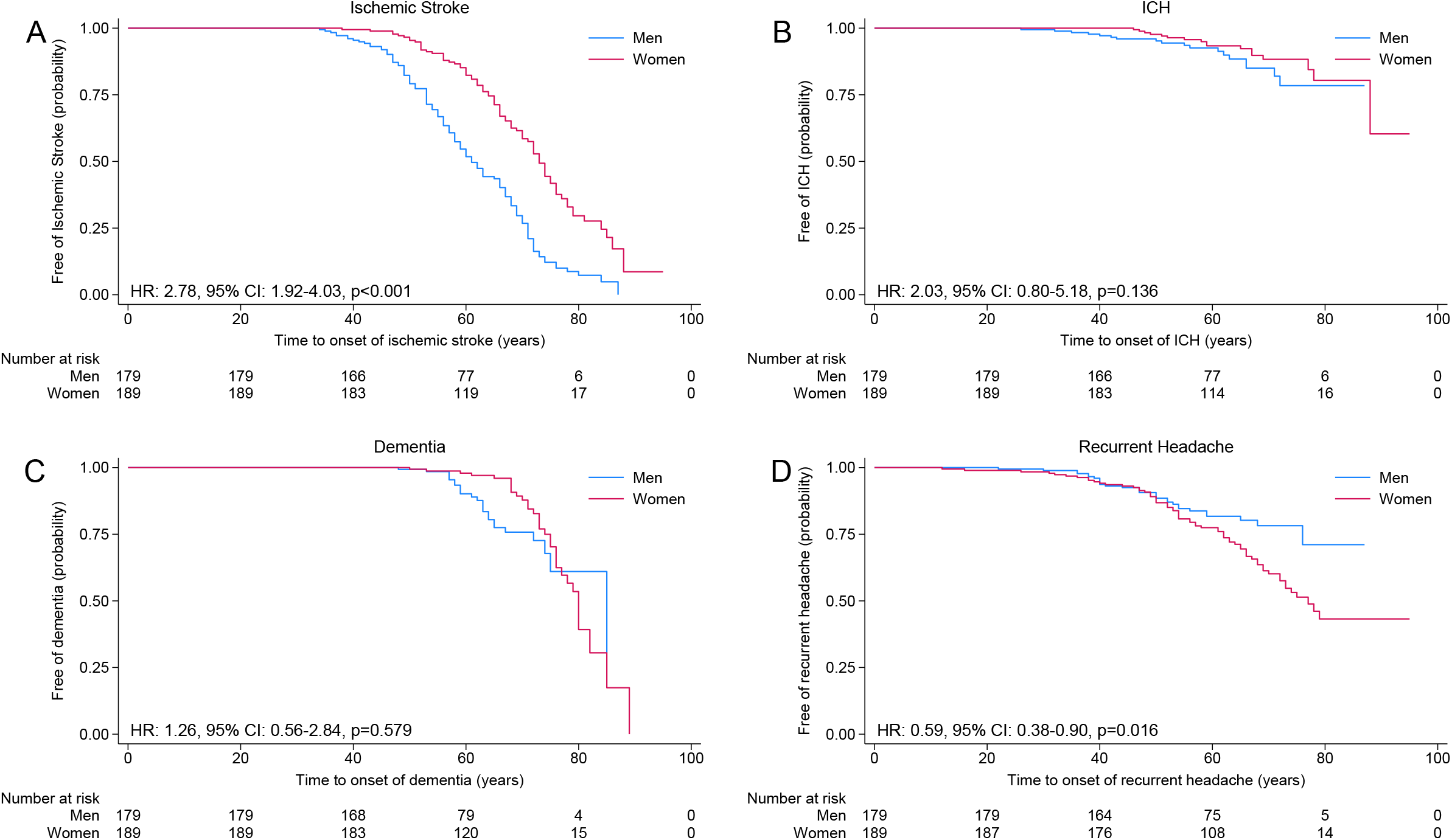
Sex differences in Kaplan-Meier estimates for the onset of ischemic stroke (A), intracerebral hemorrhage (B), dementia (C), and recurrent headache (D)

### Disability and cognitive function tests

The mRS score, K-MMSE score, and SNSB domain test results were available for 176, 174, and 96 patients, respectively. The mean age at the time of mRS and K-MMSE examinations did not differ between male and female patients (53.7 ± 3.0 years vs. 53.4 ± 3.1 years for mRS; 54.2 ± 3.0 years vs. 53.5 ± 3.0 years for K-MMSE). However, the mean age at the SNSB examination was significantly greater for male patients compared to female patients (54.7 ± 2.8 years vs. 53.5 ± 2.9 years, p=0.020). There were no significant sex differences in the mRS or K-MMSE score. Of five cognitive domains assessed with the SNSB test, only the z-scores of frontal/executive functions differed significantly between male and female (median with interquartile range, −1.81[−4.14 to −0.56] vs. −1.28[−2.54 to −0.06], p=0.022).

### Neuroradiological features

Brain MRIs were available for 304 patients. Regarding neuroradiological features, lacunes were present in 74.0% of patients, with 22.0% having more than 10 lacunes. CMBs were observed in 63.9% of patients, with 24.2% showing >10 (T2*)/>20 (SWI) CMBs. WMH in the anterior temporal region was found in 56.6% of patients, while WMH in the external capsule was observed in 89.1% of patients. The overall nWMHv was 3.9 ± 2.2% of total brain volume (Table 3). Mean age at brain MRI was 53.5 ± 3.2 years and no difference was seen between men and women (53.5 ± 3.1 years vs. 53.5 ± 3.2 years, p=0.944). Of various MRI features, men exhibited a significantly greater burden of lacunes compared to women (p<0.001) while women showed a significantly greater nWMHv compared with men (3.6±2.1% vs. 4.2±2.3%, p=0.012) in unadjusted analysis. Male sex was associated with significantly increased risk for higher category of lacune (OR 3.03, 95% CI 1.70-5.38, p<0.001) along with age (OR 1.03, 95% CI 1.01-1.05, p=0.002) and hypertension (OR 1.86, 95% CI 1.15-3.03, p=0.002) in ordinal logistic regression analysis. However, female sex was not associated with greater nWMHv compared with men after adjusting for age, variant position, and vascular risk factors (Figure 5). The sensitivity analysis with the second set of brain MRI that contained scans from 332 patients showed similar results (Figure S3 & Table S2)

**Table 3.**
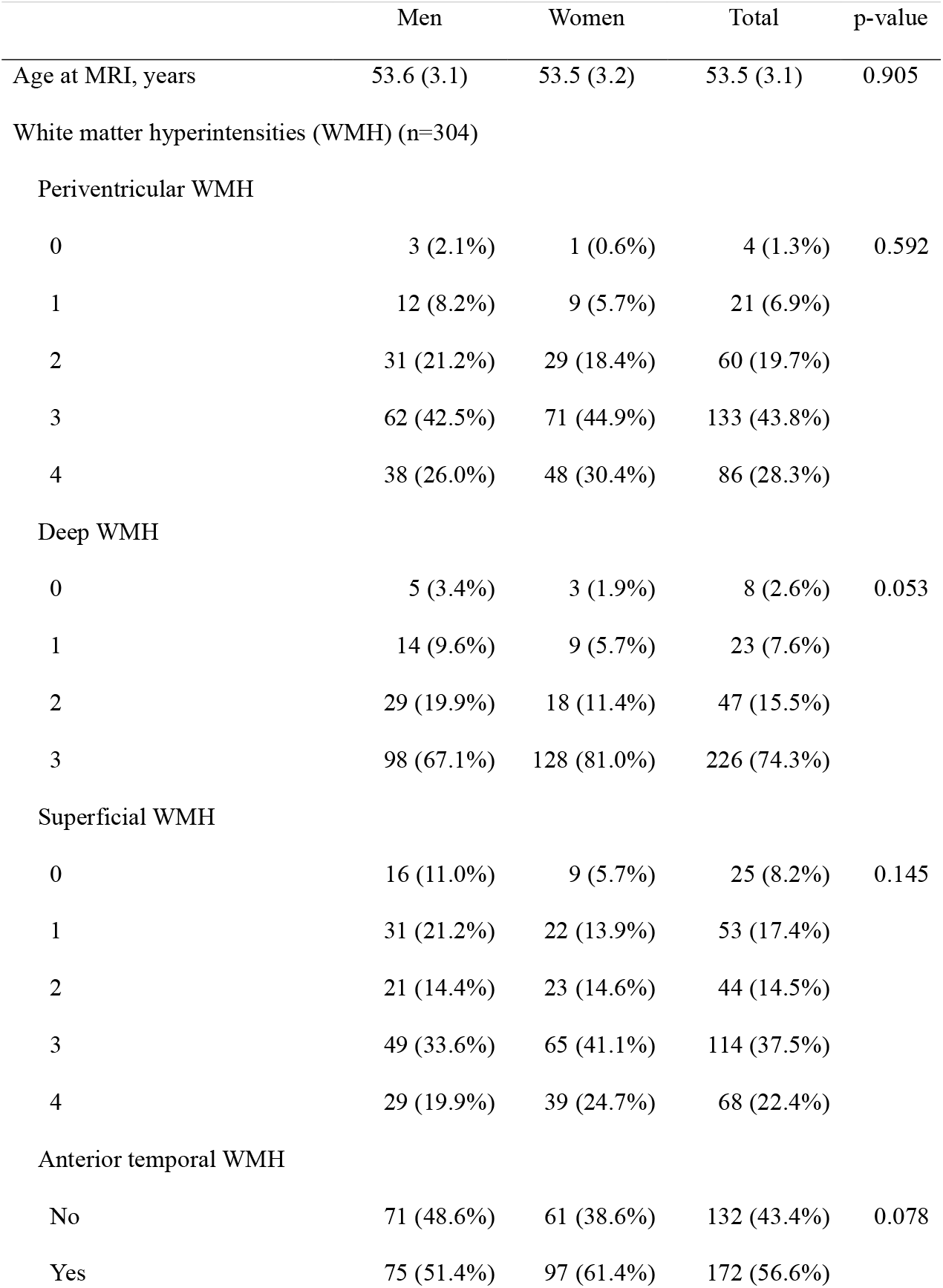
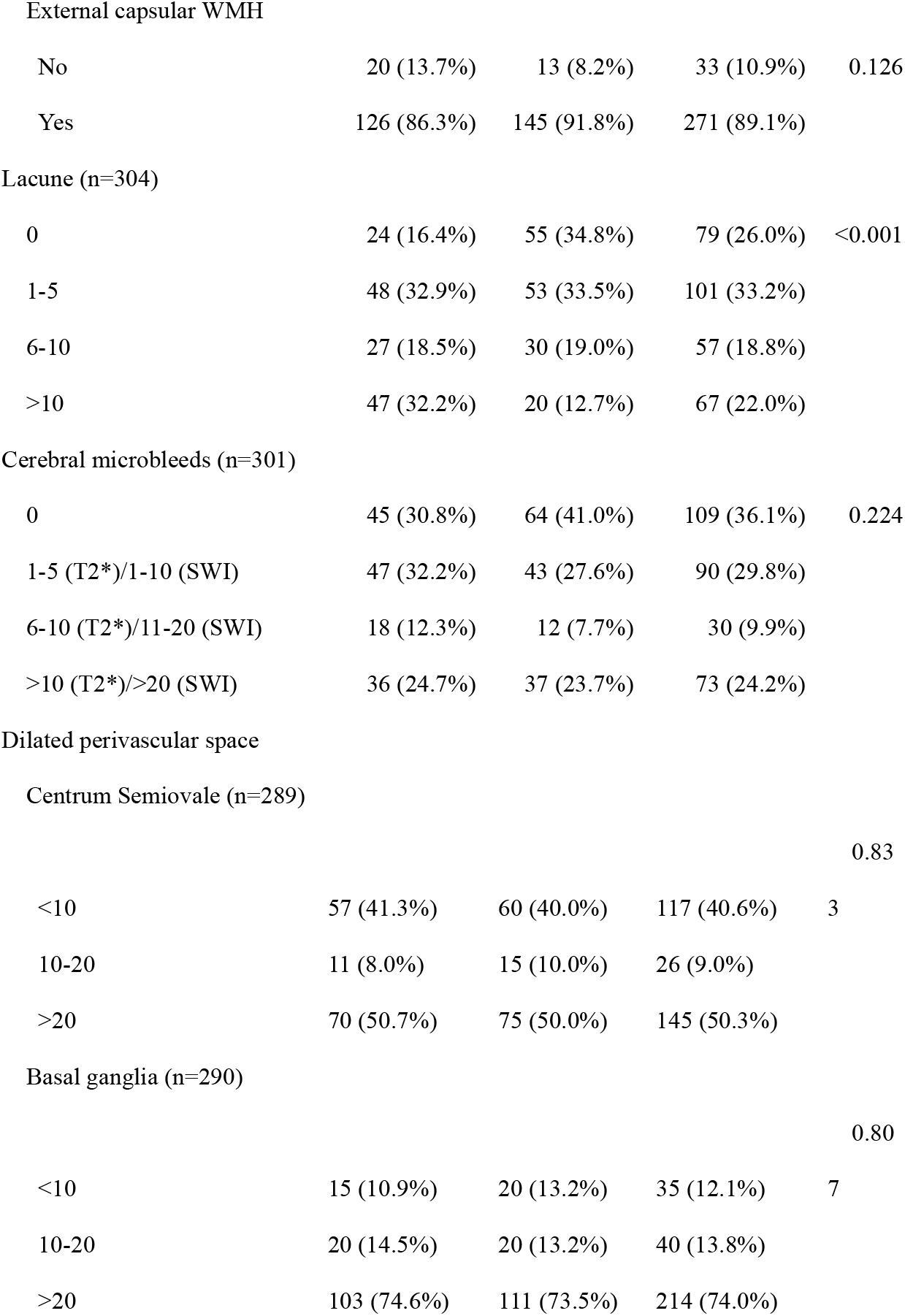
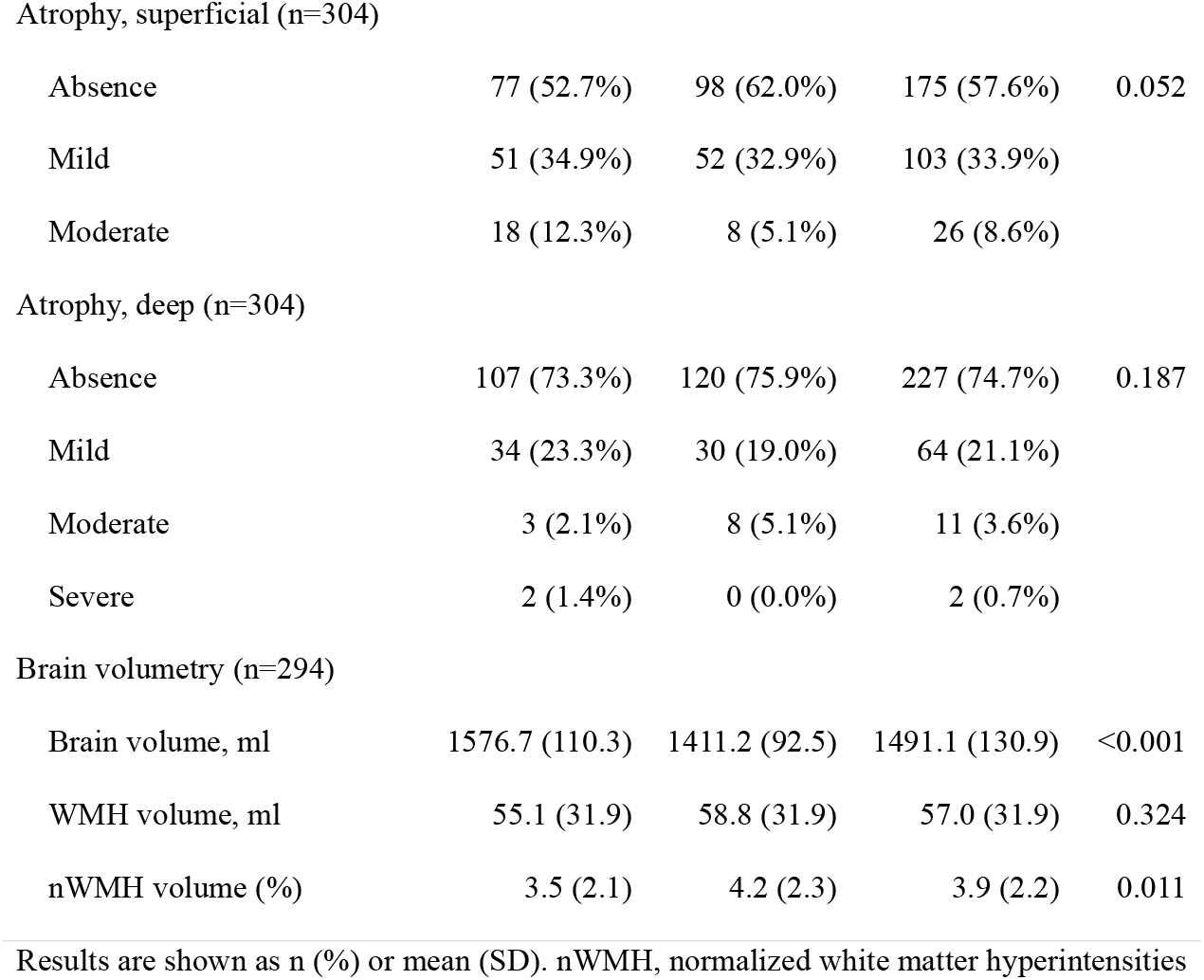
Brain MRI Characteristics of patients by sex.

|  | Men | Women | Total | p-value |
| --- | --- | --- | --- | --- |
| Age at MRI, years | 53.6 (3.1) | 53.5 (3.2) | 53.5 (3.1) | 0.905 |
| White matter hyperintensities (WMH) (n=304) |  |  |  |  |
| Periventricular WMH |  |  |  |  |
| 0 | 3 (2.1%) | 1 (0.6%) | 4 (1.3%) | 0.592 |
| 1 | 12 (8.2%) | 9 (5.7%) | 21 (6.9%) |  |
| 2 | 31 (21.2%) | 29 (18.4%) | 60 (19.7%) |  |
| 3 | 62 (42.5%) | 71 (44.9%) | 133 (43.8%) |  |
| 4 | 38 (26.0%) | 48 (30.4%) | 86 (28.3%) |  |
| Deep WMH |  |  |  |  |
| 0 | 5 (3.4%) | 3 (1.9%) | 8 (2.6%) | 0.053 |
| 1 | 14 (9.6%) | 9 (5.7%) | 23 (7.6%) |  |
| 2 | 29 (19.9%) | 18 (11.4%) | 47 (15.5%) |  |
| 3 | 98 (67.1%) | 128 (81.0%) | 226 (74.3%) |  |
| Superficial WMH |  |  |  |  |
| 0 | 16 (11.0%) | 9 (5.7%) | 25 (8.2%) | 0.145 |
| 1 | 31 (21.2%) | 22 (13.9%) | 53 (17.4%) |  |
| 2 | 21 (14.4%) | 23 (14.6%) | 44 (14.5%) |  |
| 3 | 49 (33.6%) | 65 (41.1%) | 114 (37.5%) |  |
| 4 | 29 (19.9%) | 39 (24.7%) | 68 (22.4%) |  |
| Anterior temporal WMH |  |  |  |  |
| No | 71 (48.6%) | 61 (38.6%) | 132 (43.4%) | 0.078 |
| Yes | 75 (51.4%) | 97 (61.4%) | 172 (56.6%) |  |
| External capsular WMH |  |  |  |  |
| No | 20 (13.7%) | 13 (8.2%) | 33 (10.9%) | 0.126 |
| Yes | 126 (86.3%) | 145 (91.8%) | 271 (89.1%) |  |
| Lacune (n=304) |  |  |  |  |
| 0 | 24 (16.4%) | 55 (34.8%) | 79 (26.0%) | <0.001 |
| 1-5 | 48 (32.9%) | 53 (33.5%) | 101 (33.2%) |  |
| 6-10 | 27 (18.5%) | 30 (19.0%) | 57 (18.8%) |  |
| >10 | 47 (32.2%) | 20 (12.7%) | 67 (22.0%) |  |
| Cerebral microbleeds (n=301) |  |  |  |  |
| 0 | 45 (30.8%) | 64 (41.0%) | 109 (36.1%) | 0.224 |
| 1-5 (T2*)/1-10 (SWI) | 47 (32.2%) | 43 (27.6%) | 90 (29.8%) |  |
| 6-10 (T2*)/11-20 (SWI) | 18 (12.3%) | 12 (7.7%) | 30 (9.9%) |  |
| >10 (T2*)/>20 (SWI) | 36 (24.7%) | 37 (23.7%) | 73 (24.2%) |  |
| Dilated perivascular space |  |  |  |  |
| Centrum Semiovale (n=289) |  |  |  |  |
|  |  |  |  | 0.83 |
| <10 | 57 (41.3%) | 60 (40.0%) | 117 (40.6%) | 3 |
| 10-20 | 11 (8.0%) | 15 (10.0%) | 26 (9.0%) |  |
| >20 | 70 (50.7%) | 75 (50.0%) | 145 (50.3%) |  |
| Basal ganglia (n=290) |  |  |  |  |
|  |  |  |  | 0.80 |
| <10 | 15 (10.9%) | 20 (13.2%) | 35 (12.1%) | 7 |
| 10-20 | 20 (14.5%) | 20 (13.2%) | 40 (13.8%) |  |
| >20 | 103 (74.6%) | 111 (73.5%) | 214 (74.0%) |  |
| Atrophy, superficial (n=304) |  |  |  |  |
| Absence | 77 (52.7%) | 98 (62.0%) | 175 (57.6%) | 0.052 |
| Mild | 51 (34.9%) | 52 (32.9%) | 103 (33.9%) |  |
| Moderate | 18 (12.3%) | 8 (5.1%) | 26 (8.6%) |  |
| Atrophy, deep (n=304) |  |  |  |  |
| Absence | 107 (73.3%) | 120 (75.9%) | 227 (74.7%) | 0.187 |
| Mild | 34 (23.3%) | 30 (19.0%) | 64 (21.1%) |  |
| Moderate | 3 (2.1%) | 8 (5.1%) | 11 (3.6%) |  |
| Severe | 2 (1.4%) | 0 (0.0%) | 2 (0.7%) |  |
| Brain volumetry (n=294) |  |  |  |  |
| Brain volume, ml | 1576.7 (110.3) | 1411.2 (92.5) | 1491.1 (130.9) | <0.001 |
| WMH volume, ml | 55.1 (31.9) | 58.8 (31.9) | 57.0 (31.9) | 0.324 |
| nWMH volume (%) | 3.5 (2.1) | 4.2 (2.3) | 3.9 (2.2) | 0.011 |

Results are shown as n (%) or mean (SD). nWMH, normalized white matter hyperintensities

**Figure 5.**
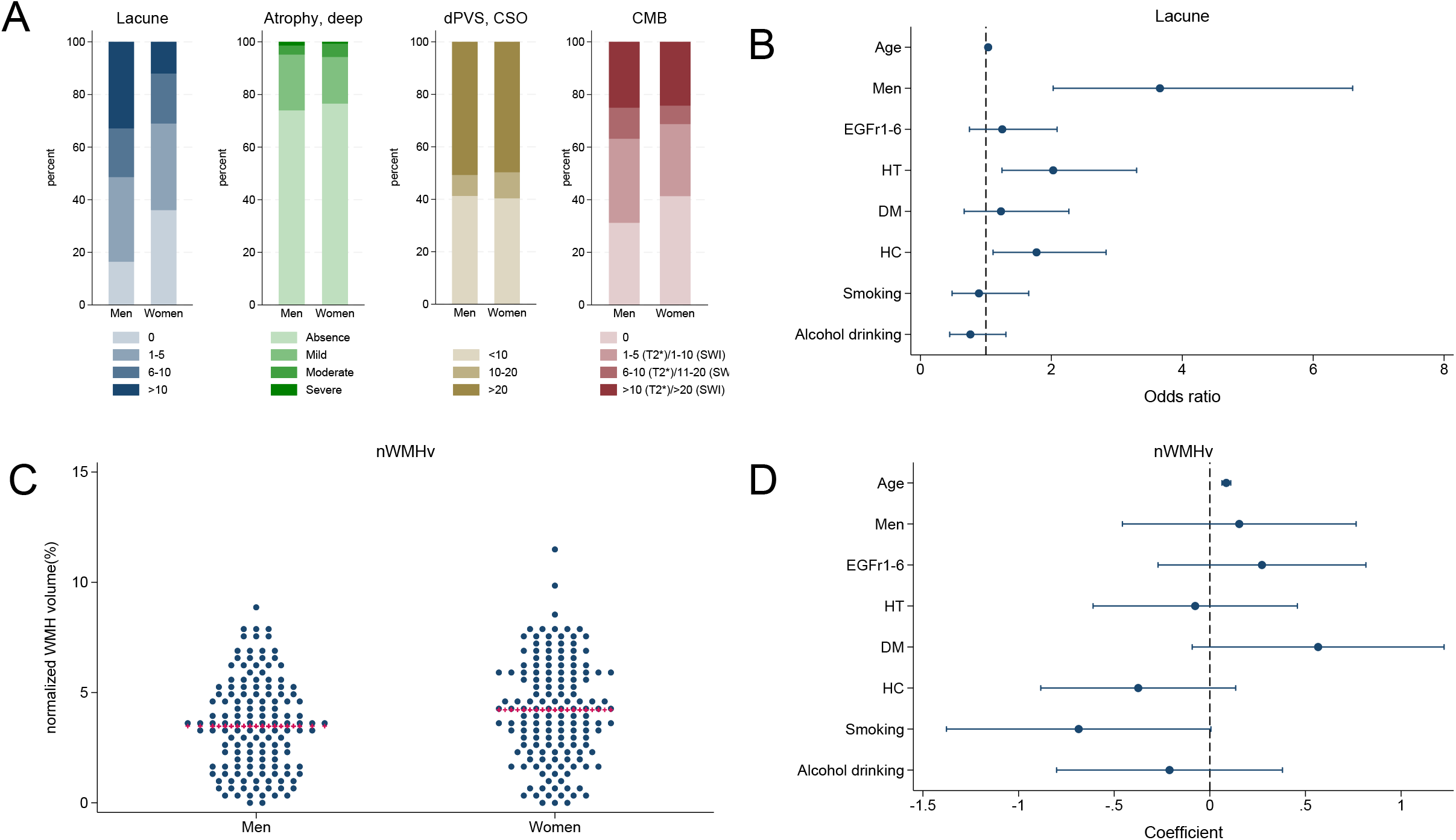
Sex differences in cerebral small vessel disease markers on brain MRI findings. (A) Distribution of lacunes, deep brain atrophy, dilated perivascular spaces (dPVS) in the centrum semiovale (CSO) and cerebral microbleeds (CMB) by sex, (B) Odds ratios for the severity of lacunes, (C) distribution of white matter hyperintensities (WMH) volume percentages by sex, horizontal red line indicate median value (D) Odds ratios for the WMH volume percentages. HT, hypertension; DM, diabetes mellitus; HC, hypercholesterolemia.

## Discussion

This nationwide study of Korean CADASIL patients revealed significant sex differences in clinical and neuroimaging features. Men exhibited a higher prevalence of ischemic stroke and an earlier onset compared to women, while women were more likely to experience recurrent headaches. The association between male sex and ischemic strokes was independent of important vascular risk factors such as hypertension or smoking. No significant sex differences were observed in the onset of ICH or dementia. Males tend to show worse frontal/executive functions compared with female on comprehensive neuropsychological assessment. On brain MRI, male sex was independently associated with an increased risk of lacunes, even after adjusting for age and vascular risk factors. Conversely, although women had a greater WMH burden in unadjusted analyses, this difference disappeared after controlling for confounders.

Our findings align with prior European studies suggesting that men with CADASIL have a higher risk of ischemic stroke and increased lacunes on brain MRI compared with women.^4,5,9^ However, some studies did not find significant sex differences in stroke onset, while others reported a trend toward earlier ischemic events in men, consistent with our results.^5,6,8^ Notably, the higher risk of ischemic stroke and its earlier onset in men could not be explained by concurrent hypertension or smoking as the risk was greater in the subgroups without hypertension or smoking. Additionally, our study confirms the previously observed association between female sex and a higher prevalence of recurrent headaches.^5^ These findings reinforce the importance of considering sex as a biological variable in CADASIL prognosis and management.

Sex differences in cerebral small vessel disease (CSVD) have been increasingly recognized, with distinct patterns in onset age and imaging features. Men tend to experience CSVD-related ischemic events, including lacunar stroke, at younger ages, likely driven by earlier and higher burden of vascular risk factors such as hypertension and smoking.^24-26^ In contrast, women often show greater burden of WMH in late life, potentially due to loss of estrogen-mediated vascular protection after menopause.^27,28^ Neuroimaging studies have shown that men have a higher prevalence of lacunes and CMBs at a given age^25^, whereas women, despite initially lower lesion burden, may exhibit faster progression of WMH and CMBs once CSVD is established.^29,30^ In CADASIL, our findings reflect this sex-specific pattern: male patients exhibited earlier onset of ischemic stroke and more lacunes, while female patients showed greater WMH volume in unadjusted analysis. These observations highlight the role of sex in modulating both the clinical expression and radiological burden of CSVD, suggesting underlying differences in pathophysiology, hormonal milieu, and vulnerability to vascular injury.^27,31^ Considering sex as a biological variable is essential for improving precision in diagnosis, risk stratification, and management strategies in CSVD, including monogenic forms such as CADASIL.

Compared to Western populations, where CADASIL-related dementia appears more common in men, our study found no significant sex difference in dementia risk even though male sex was associated with earlier and greater risk of ischemic stroke.^4,9^ It is noteworthy that our patients showed almost 20 years later onset of dementia (median age of onset 73 years) compared with the onset of dementia reported in European cohorts (50s).^4,9,32^ Previous studies have suggested that Alzheimer’s pathology could play a role in dementia among Korean CADASIL patients, which could account for the later onset of dementia and the lack of sex-based differences observed in our study.^33,34^

Additionally, no significant sex differences were observed for the prevalence or onset age of ICH in this study, although there was a trend toward earlier onset in men. Well-known risk factors for ICH in CADASIL include a history of hypertension, Asian ethnicity, and a greater burden of CMBs on brain MRI. However, sex differences have not been established as a significant risk factor.^35,36^ These findings suggest that the pathophysiological mechanisms leading to ICH in CADASIL may differ from those that lead to ischemic stroke, potentially involving distinct vascular pathologies.

Despite its strengths, this study has several limitations. First, as a retrospective analysis, it is subject to selection bias and potential inconsistencies in data collection across multiple centers. Second, while brain MRI evaluations were standardized, differences in MRI protocols among institutions may have influenced quantitative imaging measurements. Third, although we adjusted for major vascular risk factors, unmeasured confounders, such as hormonal influences and lifestyle factors, might have contributed to the observed sex differences. Fourth, cognitive assessments were not detailed in this study due to limited number of patients being examined in this retrospective study. This limits our ability to fully explore sex differences in cognitive impairment in patients with CADASIL. Finally, while this study provides important insights into a Korean CADASIL cohort, the findings may not be fully generalizable to other ethnic groups. Future prospective studies with larger sample sizes and longitudinal follow-ups are needed to further validate these findings and explore biological mechanisms underlying sex differences.

In conclusion, this study is one of the largest to examine sex differences in CADASIL within an Asian population, highlighting potential ethnic variations in disease expression. By utilizing a large, genetically confirmed cohort with standardized imaging assessments, this study advances our understanding of sex-specific disease mechanisms, which could have implications for targeted interventions and future research on sex-based therapeutic strategies.

## Data Availability

The datasets used and/or analysed during the current study available from the corresponding author on reasonable request.

## Acknowledgments

none

---

Figure S1. Factors associated with ischemic stroke stratified by hypertension and smoking status. DM, diabetes mellitus; HC, hypercholesterolemia.

Figure S2. Effect of male sex on the onset age of ischemic stroke by hypertension status. (A) Kaplan-Meier plot by sex in patients with hypertension, (B) Hazard ratios for onset of ischemic stroke in patients with hypertension (C) Kaplan-Meier plot by sex in patients without hypertension, (D) Hazard ratios for onset of ischemic stroke in patients without hypertension. DM, diabetes mellitus; HC, hypercholesterolemia.

Figure S3. Sex differences in cerebral small vessel disease markers on brain MRI findings. (A) Distribution of lacunes, deep brain atrophy, dilated perivascular spaces (dPVS) in the centrum semiovale (CSO) and cerebral microbleeds (CMB) by sex, (B) Odds ratios for the severity of lacunes, (C) distribution of white matter hyperintensities (WMH) volume percentages by sex, horizontal red line indicate median value (D) Odds ratios for the WMH volume percentages. HT, hypertension; DM, diabetes mellitus; HC, hypercholesterolemia.

Table S1. Frequency of NOTCH3 pathogenic variants

Table S2. Brain MRI findings from the second MRI scan dataset

